# A prospect on the use of antiviral drugs to control local outbreaks of COVID-19

**DOI:** 10.1101/2020.03.19.20038182

**Authors:** Andrea Torneri, Pieter Libin, Joris Vanderlocht, Anne-Mieke Vandamme, Johan Neyts, Niel Hens

**Affiliations:** Centre for Health Economic Research and Modelling Infectious Diseases, University of Antwerp, Antwerp, Belgium; Interuniversity Institute of Biostatistics and statistical Bioinformatics, Data Science Institute, Hasselt University, Hasselt, Belgium; Artificial Intelligence lab, Department of computer science, Vrije Universiteit Brussel, Brussels, Belgium; KU Leuven - University of Leuven, Department of Microbiology and Immunology Rega Institute for Medical Research, Clinical and epidemiological virology, Leuven, Belgium; Center for Global Health and Tropical Medicine, Unidade de Microbiologia, Instituto de Higiene e Medicina Tropical, Universidade Nova de Lisboa, Lisbon, Portugal

## Abstract

**Background:** Current outbreaks of COVID-19 are threatening the health care systems of several countries around the world. Control measures, based on isolation and quarantine, have been shown to decrease and delay the burden of the ongoing epidemic. With respect to the ongoing COVID-19 epidemic, recent modelling work shows that this intervention technique may be inadequate to control local outbreaks, even when perfect isolation is assumed. Furthermore, the effect of infectiousness prior to symptom onset combined with a significant proportion of asymptomatic infectees further complicates the use of contact tracing. Antivirals, which decrease the viral load and reduce the infectiousness, could be integrated in the control measures in order to augment the feasibility of controlling the epidemic.

**Methods:** Using a simulation-based model of viral transmission we tested the efficacy of different intervention measures for the control of COVID-19. For individuals that were identified through contact tracing, we evaluate two procedures: monitoring individuals for symptoms onset and testing of individuals. Moreover, we investigate the effect of a potent antiviral compound on the contact tracing process.

**Findings:** The use of an antiviral drug, in combination with contact tracing, quarantine and isolation, results in a significant decrease of the final size, the peak incidence, and increases the probability that the outbreak will fade out.

**Interpretation:** For an infectious disease in which presymptomatic infections are plausible, an intervention measure based on contact tracing performs better when realized together with testing instead of monitoring, provided that the test is able to detect infections during the incubation period. In addition, in all tested scenarios, the model highlights the benefits of the administration of an antiviral drug in addition to quarantine, isolation and contact tracing. The resulting control measure, could be an effective strategy to control local and re-emerging outbreaks of COVID-19.

## 1 Introduction

To control local outbreaks of COVID-19, we investigate the use of contact tracing and isolation in combination with an antiviral compound. Even when perfect isolation is in place, it may not be sufficient to contain a local COVID-19 outbreak. ^1^ Therefore, in the absence of a vaccine, an antiviral drug in addition to isolation could be used to contain the current COVID-19 epidemic. Today there are no Corona-specific drugs and the development of potent and safe drugs typically takes years. However, there are a number of drugs, originally targeted towards other viral infections, in clinical trials for their ability to control SARS-CoV-2 infection. Most of these drugs inhibit key components of the coronavirus infection life cycle including viral entry, replication, RNA synthesis and protein synthesis. We therefore assume that an antiviral compound will reduce the viral load of an infected individual with COVID-19. For our modeling experiments we searched for an experimental drug for which viral load data were available to inform our model. Remdesivir, was recently shown to inhibit SARS-CoV-2 in vitro^2^ and is currently under evaluation in clinical trials. Remdesivir is an investigational broad-spectrum antiviral agent that has been developed against the Ebola-virus. It functions as a nucleotide-analog inhibitor of the viral RNA-dependent RNA polymerase and has activity against a wide range of RNA-viruses. ^3^ In addition, it is also active against SARS-CoV and MERS-CoV, which is expected given the high level of similarity between the genes that are involved in the replication cycle of these corona viruses. For the aforementioned reasons we inform our model with data on the control of MERS-CoV viral load by Remdesivir in a translational murine model. ^4^ The animal model that was utilized was specifically developed to better approximate the pharmacokinetics and drug exposure profile in humans. Therefore, the measure of viral titers in lung tissue at different time-points in this model serves as a reasonable proxy for viral dynamics upon compound exposure in the controlled setting of a viral challenge. To this end, we calibrate the model to represent the viral load decrease thereof.

In this manuscript we first present the effect of isolation, considering both home quarantine (for individuals that are part of a contact trace network and for infected individuals with mild symptoms) and hospital isolation (for severe cases). We argue that when an individual is quarantined at home, this will only result in a partial reduction of contacts, accounting for contacts with household members and other isolation imperfectness. To compensate for this imperfect isolation, we consider the use of an antiviral compound. We test these different control measures in a simulation study that aims at representing, given the available information, the current COVID-19 epidemic. While at this phase of the epidemic, many countries are already beyond the point of local containment. We do however expect that the methodology we propose will be key to avoid a second peak, especially given the limited depletion of susceptibles.

## 2 Methods

### 2.1 Epidemiological model

The disease dynamics are depicted in the left panel of Figure 1. The possible transitions between epidemic classes are described by the arrows.

**Figure 1:**
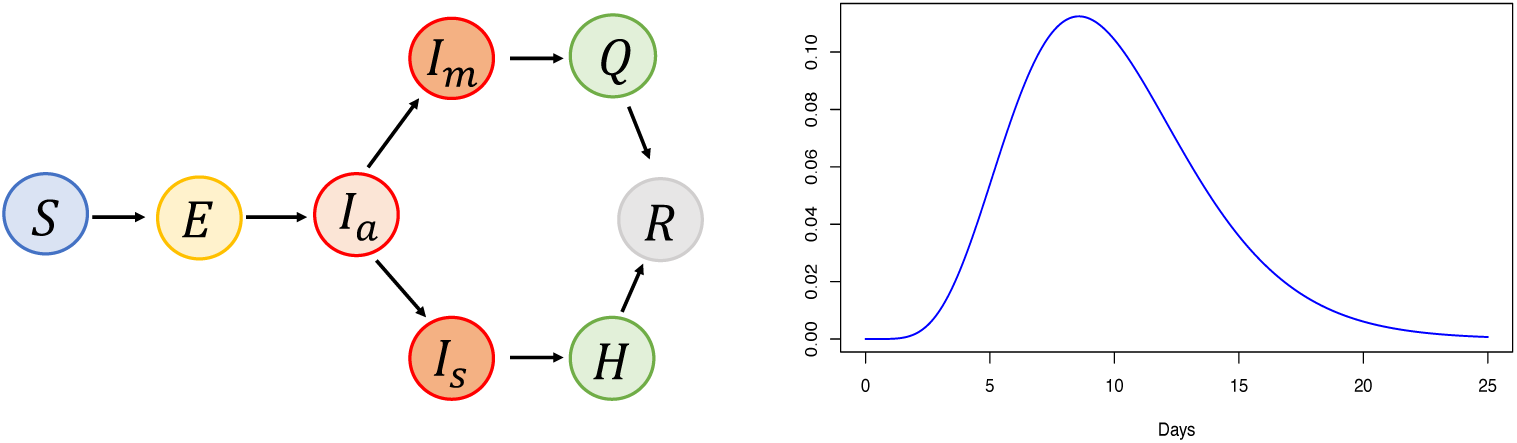
Disease dynamics: Possible transitions among the different epidemic compartments (left panel). A measure describing how infectiousness is distributed over time since infection (right panel).

Individuals are initially susceptible (*S*) and once infected, they enter the exposed class (*E*). The infection, that is at first asymptomatic (*I*_*a*_), can lead to the onset of mild (*I*_*m*_) or severe symptoms (*I*_*s*_). Symptomatic individuals are hospitalized (*H*), where they are isolated, or are confined in home quarantine (*Q*), based on the severity of symptoms. Ultimately, all infectives are assumed to either recover from infection or die (*R*). Individuals that are hospitalized are immediately isolated; therefore they can no longer transmit the disease. The quarantined individuals, can still make contacts, although at a decreased rate.

The transition from the susceptible to the exposed class is governed by a stochastic process based on the notion of infectious contact processes. ^5^ First, contacts between individuals are generated. When such contacts are generated between susceptible and infectious people, these can result in an infection event according to a Bernoulli probability value based on the time since infection. This probability is computed, at a precise time point, as the product of two components: the infectiousness measure, *ν*(*t*), which quantifies the level of infectiousness over time, and the total amount of infectivity *q*, i.e. the number of infections over the contact rate.^6^ The function *ν*(*t*) is defined over the exposed and infectious period, or analogously over the incubation and symptomatic period, along which it integrates to one. This function is scaled to have a similar shape among different infectives, based on their lengths of exposed and infectious period. According to this framework, an infectious individual makes effective contacts at a rate, *r*(*t*) given by:

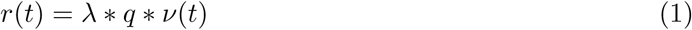

where *λ* is the contact rate. The mean number of effective contacts is an approximation of the reproduction number. The two quantities are identical in an infinite and homogeneous population, where the probability of making two effective contacts with the same person is zero. For the considered population size, the probability of this event is extremely low. Therefore, throughout the manuscript, we approximate the reproduction number with the mean number of secondary cases.

In the considered framework, isolation/quarantine is implemented by reducing the contact rate *λ* at the time of diagnosis. To date, little is known about the difference in viral load among severe and mild cases. Zhou et al. ^7^ indicate that in nasal and throat swabs the viral load is higher in mild cases. Due to this uncertainty, we assume that the same curve is defined for all the infected individuals. In this work, we assume the population to be homogeneous, closed and finite population. These assumptions relate to the control measures currently in place, e.g. in Italy, aiming at containing immigration and emigration in a country with an ongoing outbreak.

### 2.2 Simulation parameters and distributions

In Table 1 we report the parameters and distributions that were utilized in the simulation study. Where distributions are not reported, the parameters are assumed to be constant. In the last column, when available, we report the references to the literature that justifies the choice of the parameter value, or distribution, we use.

**Table 1:** Model Parameters

| Name | Mean Value (sd) | Distribution | Reference |
| --- | --- | --- | --- |
| Incubation period | 5.2 days (2.8 days) | Weibull | 8,9 |
| Symptomatic period length | 18 days | Exponential | 7 |
| Time to hospitalization | 1.6 days | Exponential | 10 |
| Reproduction number: | 2.5 | NA | 8,9,11,12 |
| Symptomatic Individuals | 100% | NA | Assumed |
| Severe cases | 16% | NA | 13 |
| Population size | 500 | NA | Motivated in the text |
| Daily contact rate | 12 contacts | NA | 14 |
| Infectiousness measure | 10 days (3.8 days) | Gamma | 7,15,16 |

We assume infectious individuals to make, on average, between two and three effective contacts. This value, is set accordingly to the current estimate of the basic reproduction number. ^8^,9,11,12 The infectiousness measure is set to represent the viral load observations reported in ^7^,15,16, the peak of which is reached within a few days after symptoms onset. The total length is modeled as the convolution of incubation and symptomatic period. Incubation and symptomatic period, are set, respectively, to have a mean length of 5·2 days and 18 days. There is no precise estimate of the length of the symptomatic period to date, thus we choose it according to the aggregated nasal and throat swabs data. ^7^ The time to hospitalization is estimated from the data presented in Backer et al. ^10^ We assume that the time to hospitalization coincides with the time of diagnosis. At this time-point, depending on the severity of the symptoms, individuals are isolated (severe cases) or quarantined (mild cases). The population size is set to 500 to represent a localised outbreak of COVID-19. Furthermore, we assume that the contact tracing starts when individuals are diagnosed.

### 2.3 Contact tracing and isolation

In order to implement contact tracing we keep track of a contact history *H*_*i*_ for each individual *i* for all contacts made since the time of infection. When an individual *i* is found to be infected with SARS-CoV-2, a contact tracing procedure is started. We assume that each contact in *H*_*i*_ will be traced back successfully with probability *η*. Depending on the considered scenario, traced-back individuals will be monitored or tested. When individuals are found positive for SARS-CoV-2, they will be put in quarantine/isolation. For certain scenarios, next to quarantine, we also inject infected individuals with an antiviral drug (i.e., Remdesivir).

We assume that the test can detect infection after two days since infection. Traced individuals who test negative the first time, are tested again after two days. The quarantine will result in a decreased contact rate (i.e. imperfect isolation), *λ*_*q*_, while in case of perfect isolation the contact rate is set to zero. Similarly, diagnosed individuals will also be quarantined: at home (mild symptoms), with a decreased contact rate *λ*_*q*_, or in the hospital (severe symptoms), where we assume that perfect isolation is possible. In all the considered scenarios, the 16% of individuals is isolated while the rest is put in quarantine. The selected proportion reflects the proportion of severe cases reported in Guan et al. ^13^

### 2.4 Antiviral compounds

To compensate for imperfect isolation we investigate the use of antiviral compounds to reduce the infectiousness of an infected individual. We assume that, once the antiviral compound has been administered, the infectiousness measure will exponentially decay according to an inverse Malthusian growth model (shown in Figure 2). ^17^ The rate of this decay is set to represent the reduction in viral load, due to Remdesevir, as reported in ^4^ for the MERS coronavirus.

**Figure 2:**
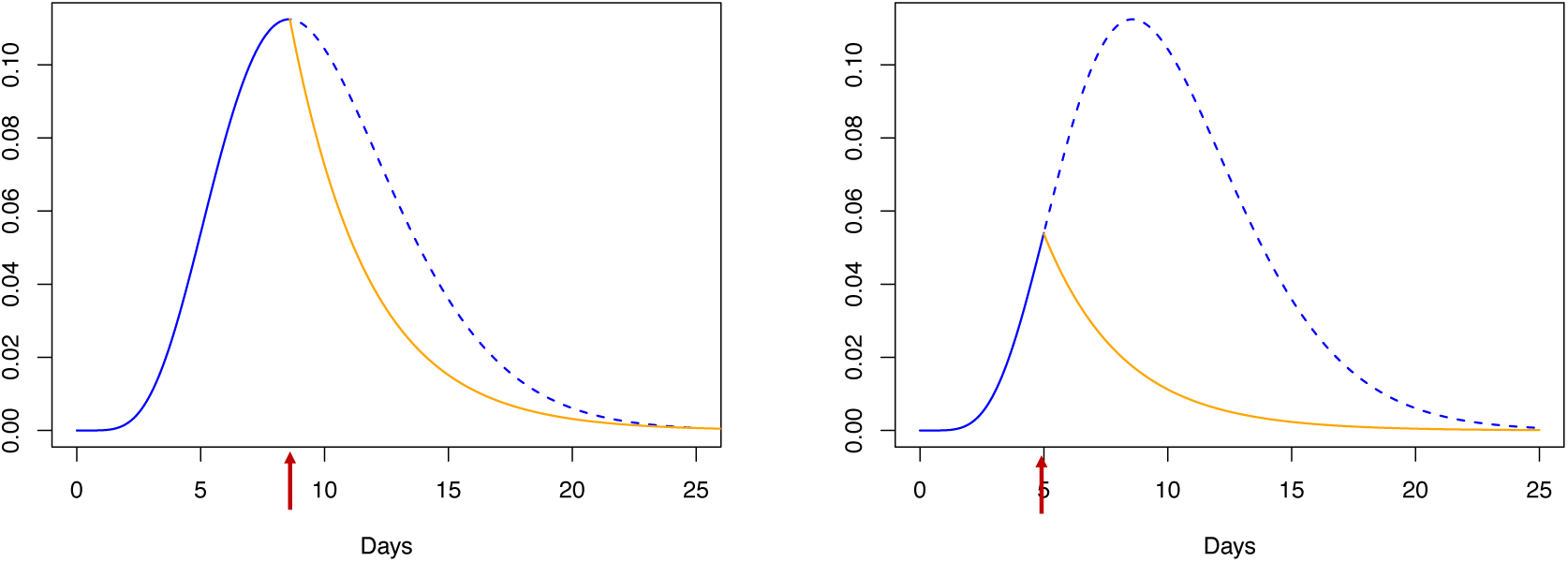
Reduction of infectiousness. The blue and the orange lines describe the infectiousness measure, respectively, before (dashed blue) and after antiviral injection (solid yellow). The red arrows indicate the injection times.

### 2.5 Scenarios

We assume the following parameters for the reduction of contacts because of home quarantine: *λ*_*q*_ = 0·1*λ*, 0·25*λ*, 0·5*λ* and for the probability of tracing back a contact in history *H*_*i*_: *η* = 0·25,0·5,0·75^18^. Scenarios: In all the considered scenarios we assume that individuals are isolated, or quarantined, when diagnosed. Moreover, we assume that contact tracing starts at the time of diagnosis.

IAS: Traced individuals are monitored for two weeks, and isolated/quarantined if they show symptoms during this period. This scenario is similar to the baseline scenario described by Hellewell et al. ^1^ with the exception that in our description only severe cases are isolated while the mild are home quarantined. This scenario reflects more realistically the current practice of containment.

IBS: Traced individuals are isolated/quarantined, as soon as they test positive for SARS-COV-2. We assume that an individual that is infected tests positive 2 days after infection. Therefore, a traced individual is tested immediately when traced, and, if this test was negative, we test the individual again two days later.

IBTBS: A diagnosed patient is immediately treated with the antiviral drug. Furthermore, traced individuals are isolated/quarantined and injected with the antiviral drug, as soon as they test positive for SARS-COV-2. We assume that an individual that is infected tests positive 2 days after infection. Therefore, a traced individual is tested immediately when traced, and, if this test was negative, we test the individual again two days later.

For each scenario we run 5000 simulations. Among these, we compute the final size and the cases at peak for the one in which at least the 10% of individuals have been infected. Doing this, we only account for outbreaks that are most challenging to contain.

## 3 Results

Quarantine, isolation and antiviral treatments lead, in different levels, to the mitigation of the outbreak by reducing the final size as well as by reducing the number of cases at the peak of the epidemic. The containment performance depends, among all the scenarios, on the probability to successfully trace contacts and on the reduction in contact rate due to quarantine (Figure 3). Isolation and quarantine lead to a substantial decrease in final size and peak incidence. When performed prior to symptom onset their efficacy increases, which is important, as recent work indicates that infectees are infectious prior to symptom onset.^19^ The antiviral treatment is shown to have a substantial impact and, together with quarantine and isolation, significantly reduces the final size, the peak incidence and the number of outbreaks that are most challenging to contain.

**Figure 3:**
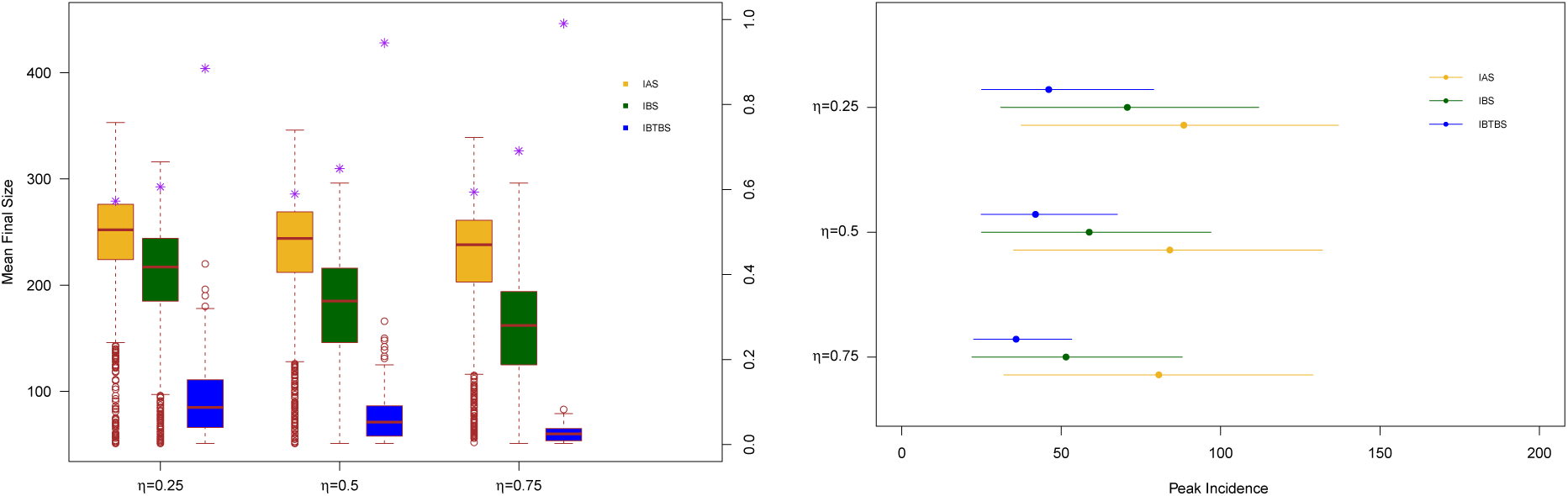
Final size and Mean peak incidence. Left panel: distributions of the final size value for Scenario IAS (yellow), Scenario IBS (green) and Scenario IBTBS (blue) when the quarantine contact rate is *λ*_*q*_ =0·25*λ* together with the probability that a simulation leads to a number of cases smaller than the 10% of the population (purple asterisks). Right panel: mean peak incidence together with the 2·5 % and the 97·5 % percentiles.

## 4 Discussion

We assume that we have sufficient antiviral drugs doses to treat all individuals that are encountered via the contact tracing procedure. This is motivated by the fact that we consider an emerging outbreak and the required number of doses will thus be limited.

Furthermore, we assume that all the individuals will show symptoms, sooner or later, during their infectious period, and therefore all infected individuals will be diagnosed. Due to the awareness of COVID-19 given by media and government officials, individuals are more likely to act upon even mild symptoms. This assumption is in line with the work of Hellewell et al. ^1^ Moreover, we assume that all infected individuals will eventually be diagnosed. While this is a limitation of our study, we argue that in the scenarios where we aggressively trace and treat the contacts of individuals, we are more likely to find (and constrain) cases that would otherwise go undetected. To challenge this assumption, we simulate the impact of asymptomatic individuals, defined to be infectives that do not show symptoms during their infectious periods. Results reported in the sensitivity analysis show that, for the tested scenarios, the use of an antiviral drug increases the control of the outbreak, even when undiagnosed individuals are circulating in the population.

In the hospital, we assume perfect isolation, meaning that infected individuals cannot spread the infection. While this assumption is in line with earlier works ^1^, health care workers are at risk and they could be infected by infected individuals in isolation.

In the IBS and IBTBS scenarios we assume that the traced individuals that test positive are isolated in the 16% of cases, even before showing actual severe symptoms.

Although this model is informed with data on the control of MERS-CoV viral load using prophylaxis with Remdesivir, it stands to reason that different classes of viral inhibitors control the viral load in different ways. Additionally, despite the sequence similarity of MERS-CoV and SARS-COV-2 it remains to be established whether the impact of Remdesivir (or other antivirals) on the viral load is similar. To this end, longitudinal data of the viral load on COVID-19 infected patients treated with different viral inhibitors will be informative. Furthermore, Sheahan et al. ^4^ demonstrated that the degree of the clinical benefit of Remdesivir for MERS depends on the viral dose and also on the timing of the treatment of the viral inhibitor. A lethal viral dose and delaying the initiation of antiviral treatment failed to fully prevent viral pathogenesis. Although Remdesivir proved effective at reducing the viral load also in these conditions, the argument for an early start of antiviral treatment is evident. Presumably, reducing the viral load with an antiviral compound loses its efficacy in advanced disease as the tissue damage is sustained by inflammatory processes in absence of the viral initiator. In our implementation, antiviral injections are immediately administered to successfully traced back individuals, mostly in their asymptomatic phase, and to the diagnosed patient. Therefore, we believe that the assumptions on the use of this drug, in the considered scenario, are reasonable.

## 5 Conclusion

The ongoing epidemic of COVID-19 threatens the health system of many countries. Although control measures such as isolation and quarantine are of great importance, relying on their exclusive use could fail to contain an outbreak. In addition, when a proportion of infected individuals requires hospital care, the number of cases at peak should be minimized as much as possible to avoid that regional health care infrastructure is overwhelmed. In the current study we highlight the impact of an antiviral compound that reduces the viral load and, consequently, the infectiousness of infectives. For this simulation study we utilized the data on the control of the viral load by Remdesivir in a pathogen challenge experiment, however, this study can be easily extended to other antivirals, given the availability of viral load data upon drug administration. Although the efficacy of administering an antiviral compound, in addition to isolation and quarantine depends on the effectiveness of the respective drug, it is plausible that drugs that interfere with the infection life cycle show a comparable impact on the viral load. We demonstrate that the implementation of such a compound together with quarantine leads to a substantial reduction of the final size and the peak incidence. In addition, the number of outbreaks that are most challenging to contain decreases when the antiviral is administered to diagnosed and traced individuals. Therefore, the administration of an antiviral drug, together with isolation and quarantine, is expected to have a major impact in the control of local COVID-19 outbreaks. Although antiviral compounds are mostly evaluated in clinical trials for their capacity to control the disease manifestations and alter the clinical outcome in individual patients, our study implies that antiviral compounds can be implemented to alter the spread of a viral epidemic by influencing the viral load and infectiousness of infectees upon quarantine. Additionally, it is expected that the implementation of such a strategy during an epidemic outbreak might also prove effective to prevent hospitalisation and the pressure on the health care capacity as these antiviral treatment is initiated earlier in the disease.

Currently, there remains a great deal of uncertainty with respect to the proportion of asymptomatic infectees. Therefore, we performed a sensitivity analysis where we consider a proportion of 35% asymptomatic individuals, which is in line with the findings of recent works. ^20,21^,For this proportion we consider a setting where the reproductive number between symptomatic and asymptomatic is equal (Figure 9) and a setting where the asymptomatic infectees have a reproduction number that is 55% of the reproduction number set for symptomatic infectees (Figure 10).^20^ For both settings, we show a significant advantage with respect to the reduction of the final size and the peak incidence. Yet, our analysis also shows that when the asymptomatic are as infectious as the symptomatic infectees, it is important to have a high contact tracing success rate (*η*).

We remain hopeful that the ongoing clinical trials will reveal an antiviral compound that can be used as a treatment and prophylaxis. Yet, our work shows that such compounds have an additional potential to mitigate the pandemic when implemented in concert with quarantine and contact tracing. Therefore research aimed at the identification of new drugs targeting different viral families with pandemic potential is warranted.

## Data Availability

Our study concerns the modelling of antiviral compounds to contain COVID-19 epidemics. Our modelling code has been posted to github.

https://github.com/AndreaTorneri/ViralTransm

## Acknowledgements

This work is funded by the EpiPose project from the European Union’s SC1-PHE-CORONAVIRUS-2020 programme, project number 101003688 and by the European Union’s Horizon 2020 research and innovation programme (grant agreement 682540 — TransMID). A.T. acknowledges support from the special research fund of the University of Antwerp. P.L. was supported by funding from the Flemish Government under the “Onderzoeksprogramma Artificiële Intelligentie (AI) Vlaanderen” programme.

## Author contributions

A.T. contributed to the conceptualization of the study, the construction and implementation of the mathematical model, the experimental setting and the writing of the manuscript. P.L. contributed to the conceptualization of the study, the construction of the mathematical model, the experimental setting and the writing of the manuscript. J.V. contributed to the construction of the mathematical model, the experimental setting and the writing of the manuscript. A.V. contributed to the conceptualization of the study and the writing of the manuscript. J.N. contributed to the conceptualization of the study and the writing of the manuscript. N.H. contributed to the conceptualization of the study, the construction of the mathematical model, the experimental setting, the writing of the manuscript and the project supervision.

## Competing interests

Besides his employment at the Hasselt University, JV is employed at Bioqube Ventures. Bioqube Ventures was not involved in this work, nor does it prosper financially as a result of the current study. The other authors declare that they have no competing interests.

## Materials and Correspondence

Correspondence and material requests should be addressed to Prof. Dr. Niel Hens. Source code of our model and experiments can be found on https://github.com/AndreaTorneri/ViralTransm.

## Appendix: sensitivity analyses

We report in Figure 4 and 5 the sensitivity analysis for the quarantine contact rate: *λ*_*q*_ = 0.1*λ*, 0.5*λ*. The introduction of an antiviral compound substantially contributes in reducing the final size, the peak incidence and the probability of a challenging outbreak in all the considered settings. This decrease, compared to the scenario in which only isolation/quarantine is implemented, increases when quarantine is less effective (left panels). In Figure 6 and 7, we vary the reproduction number that is set, respectively, to *R*_0_ = 2 and *R*_0_ = 3. The effect of the antiviral drug, in addition to isolation and quarantine, increases when the reproduction number increases. In case of *R*_0_ = 3, the peak incidence decreases when the antiviral compound is used, compared to control measures based only on isolation and quarantine after symptoms onset.

**Figure 4:**
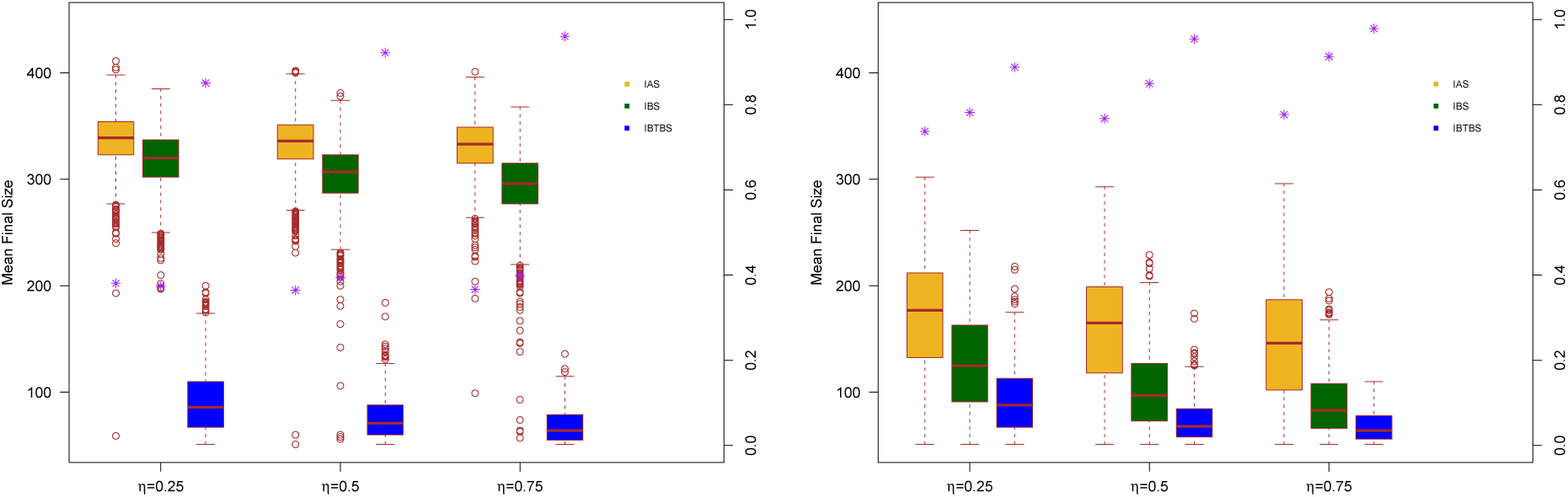
Distributions of the final size value for Scenario IAS (yellow), Scenario IBS (green) and Scenario IBTBS (blue) when the quarantine contact rate is *λ*_*q*_=0·5*λ* (left panel) and *λ*_*q*_=0·1*λ* (right panel) together with the probability that a simulation leads to a number of cases smaller than the 10% of the population (purple asterisks)

**Figure 5:**
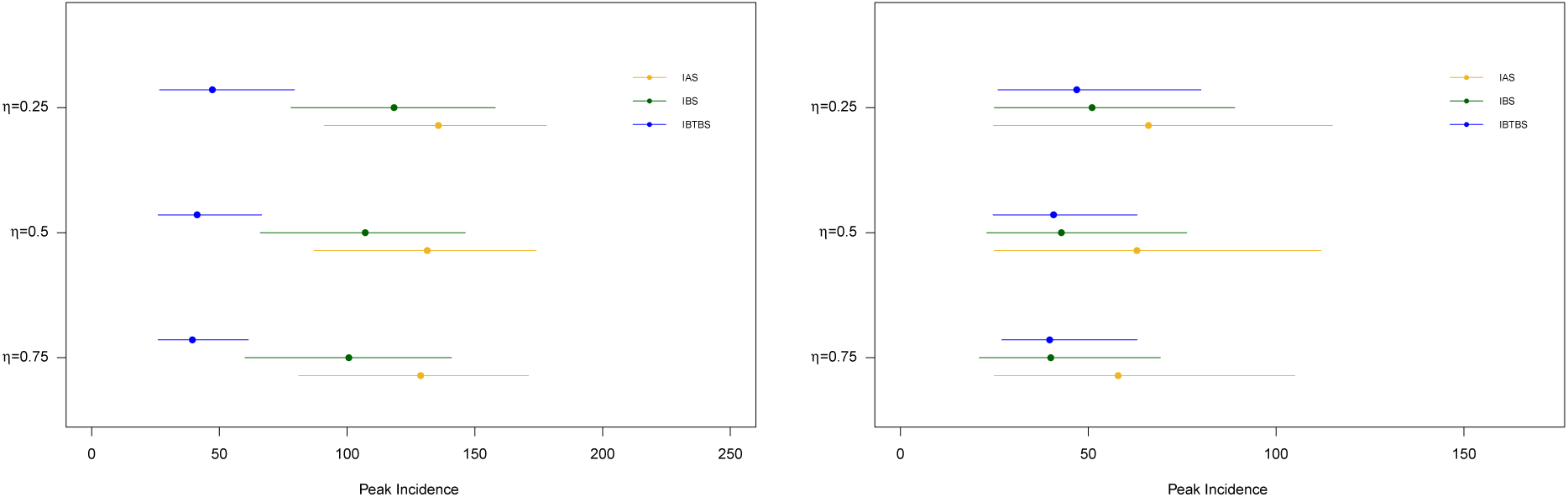
Mean peak incidence for Scenario IAS (yellow), Scenario IBS (green) and Scenario IBTBS (blue) when the quarantine contact rate is *λ*_*q*_=0·5*λ* (left panel) and *λ*_*q*_=0·1*λ* (right panel) together with 2·5% and 97·5% percentiles.

**Figure 6:**
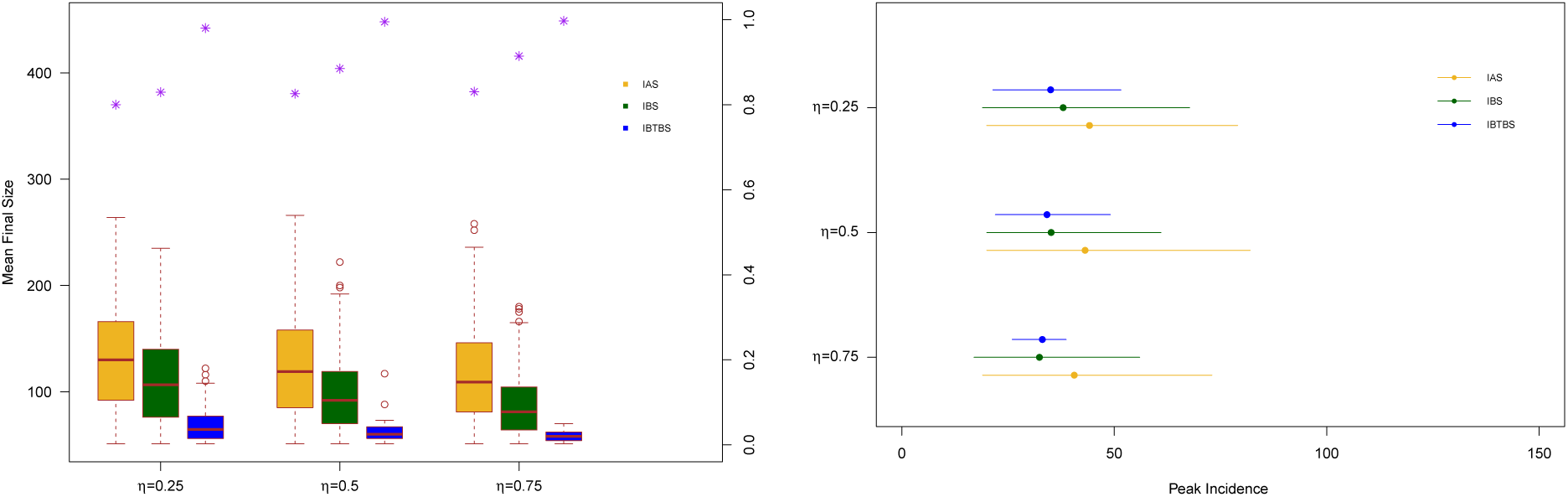
Final size distribution (left panel) and mean peak incidence for Scenario IAS (yellow), Scenario IBS (green) and Scenario IBTBS (blue) when the quarantine contact rate is *λ*_*q*_=0·25*λ*, and *R*_0_ = 2. In the left panel, for each scenario we report the probability that a simulation leads to a number of cases smaller than the 10% of the population (purple asterisks). In the right panel, together with the point estimates we report the 2·5% and 97·5 % percentiles.

**Figure 7:**
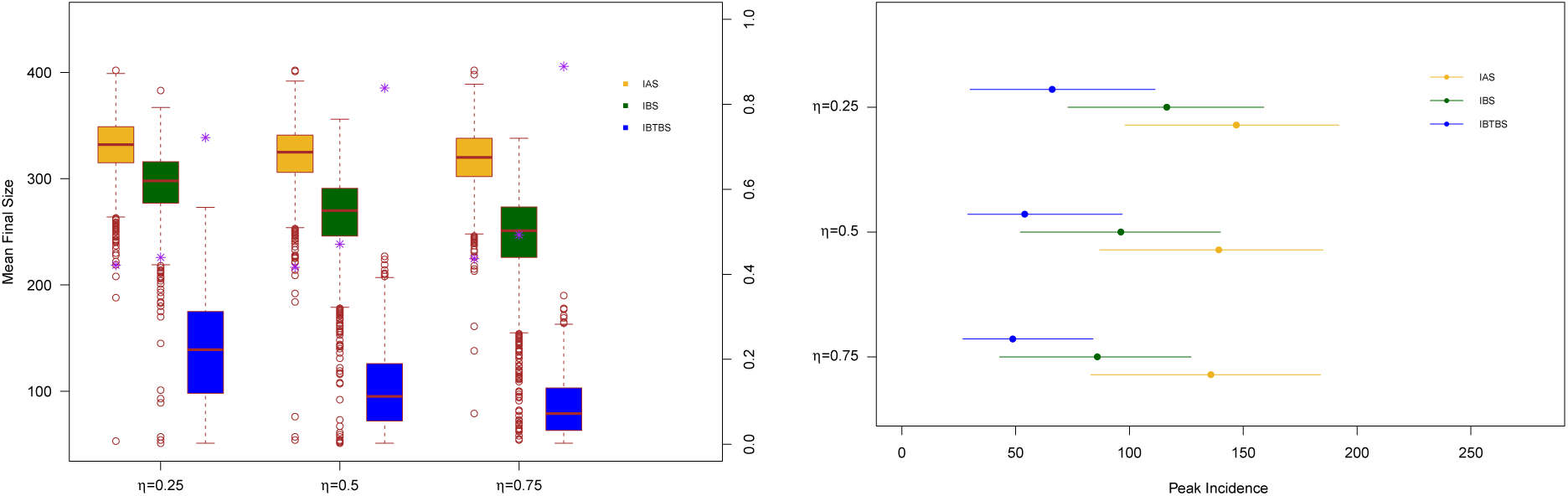
Final size distribution (left panel) and mean peak incidence for Scenario IAS (yellow), Scenario IBS (green) and Scenario IBTBS (blue) when the quarantine contact rate is *λ*_*q*_=0·25*λ*, and *R*_0_ = 3. In the left panel, for each scenario we report the probability that a simulation leads to a number of cases smaller than the 10% of the population (purple asterisks). In the right panel, together with the point estimates we report the 2·5% and 97·5 % percentiles.

In Figure 8 we investigate the effect of a longer time needed for the test to detect an infectious individual.We assume the test is positive when performed on an infectious individual who has been infected since at least 4 days. Simulations show a substantial increase, both in the final size and the peak incidence for the IBS scenario. Instead, the use of an antiviral drug results also in this case of remarkable impact in both the final size and the peak incidence.

**Figure 8:**
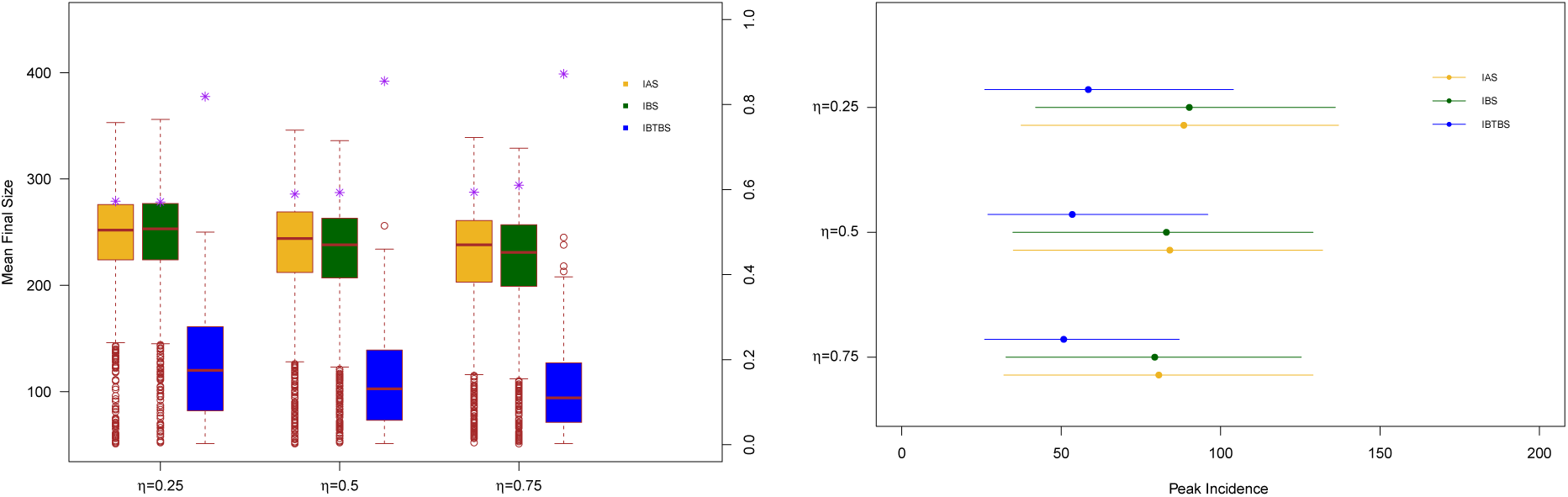
Final size distribution (left panel) and mean peak incidence for Scenario IAS (yellow), Scenario IBS (green) and Scenario IBTBS (blue) when the quarantine contact rate is *λ*_*q*_=0·25 *λ, R*_0_=2·5 and the test detect positively an infectious individual after 4 days since infection. In the left panel, for each scenario we report the probability that a simulation leads to a number of cases smaller than the 10% of the population (purple asterisks). In the right panel, together with the point estimates we report the 2·5% and 97·5 % percentiles.

We tested also the effect of an asymptomatic proportion of infectives. We assume that the 35% of individuals will not show symptoms during their infectious periods, according to result reported in recent studies. ^20^,21 We consider two values of the reproduction number for the asymptomatic individuals. In Figure 9 asymptomatic and symptomatic individuals have the same reproduction number of value *R*_0_ = 2.5, while in Figure 10 the reproduction number for symptomatic and asymptomatic is set, respectively, to 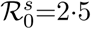 and 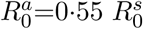. Also in these scenarios we noticed that the administration of an antiviral drug lead to a substantial decrease in the final size and peak incidence compared to the values obtained when the other control measures are considered.

**Figure 9:**
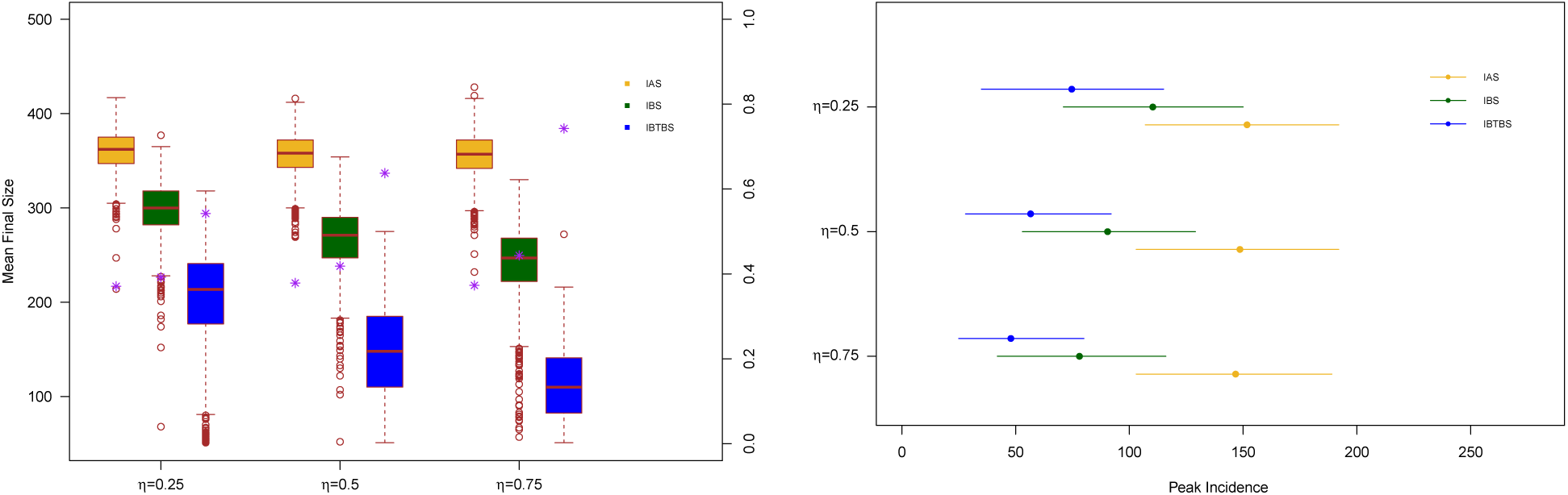
Final size distribution (left panel) and mean peak incidence for Scenario IAS (yellow), Scenario IBS (green) and Scenario IBTBS (blue) when the quarantine contact rate is *λ*_*q*_=0·25 *λ* and the proportion of asymptomatic individuals is 35%. We consider the same reproduction number for asymptomatic 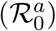.and symptomatic 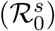 individuals: 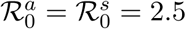In the left panel, for each scenario we report the probability that a simulation leads to a number of cases smaller than the 10% of the population (purple asterisks). In the right panel, together with the point estimates we report the 2·5% and 97·5 % percentiles.

**Figure 10:**
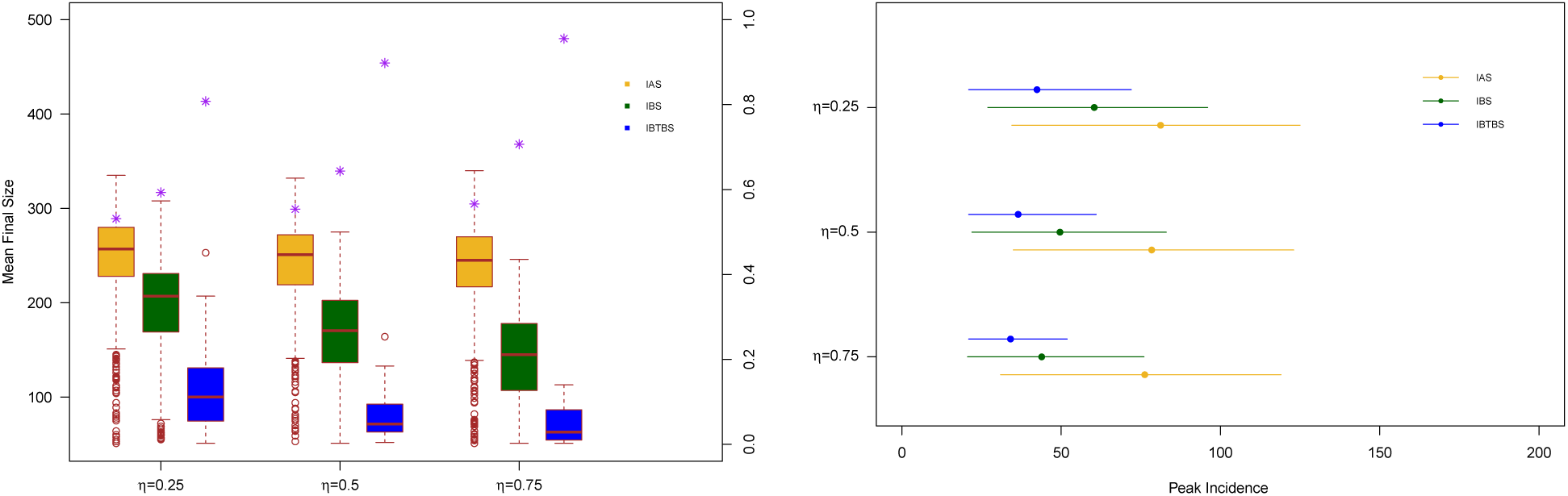
Final size distribution (left panel) and mean peak incidence for Scenario IAS (yellow), Scenario IBS (green) and Scenario IBTBS (blue) when the quarantine contact rate is *λ*_*q*_=0·25 *λ, R*_0_=2·5 and the proportion of asymptomatic individuals is 35%. We consider the a reproductive number that is different for asymptomatic 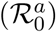 and symptomatic 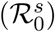 individuals: 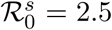 and 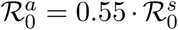 In the left panel, for each scenario we report the probability that a simulation leads to a number of cases smaller than the 10% of the population (purple asterisks). In the right panel, together with the point estimates we report the 2·5% and 97·5 % percentiles.

